# High Sensitivity of Facility-Level Wastewater Surveillance for Detecting Respiratory Virus Surges in Large Municipal Hospitals in New York City

**DOI:** 10.64898/2026.08.04.26358888

**Authors:** Sofia Pesantez, Madhura Rane, Sherin Kannoly, Leopolda Silvera, Nash Rochman, Aiden Stanciu, Valeria Martinez, Sukhleen Kaur, José Pagán, Monica Trujillo, John Dennehy, Denis Nash

**Affiliations:** Institute for Implementation Science in Population Health (ISPH), City University of New York School of Public Health, New York, NY, USA; Dr. James A. Ferguson Emerging Infectious Diseases RISE Fellowship at the Kennedy Krieger Institute; Biology Department, Queens College, City University of New York; Public & Global Health/Biosurveillance, NYC Health + Hospitals; Computational Biology Branch, Division of Intramural Research, National Library of Medicine, National Institutes of Health, Bethesda, MD, USA; Department of Epidemiology and Biostatistics, City University of New York School of Public Health, New York, NY, USA; Department of Biological Sciences and Geology, Queensborough Community College of The City University of New York, Queens, NY, 11364, USA

**Keywords:** Viral Epidemiology, Wastewater surveillance, PMMOV, Digital PCR

## Abstract

Hospital-based wastewater surveillance may complement community and clinical surveillance data in important ways, and may be useful in jurisdictions without community-based wastewater surveillance. From May 2024-April 2026, we analyzed weekly samples (n=190) from three hospitals in New York City using digital PCR to evaluate the sensitivity, specificity, and positive predictive value (PPV) of wastewater viral detection against facility SARS-CoV-2 and influenza A/B inpatient caseloads. Sensitivity was 38-42% for SARS-CoV-2 and 36-49% for influenza A/B, while specificity exceeded 72% for all pathogens. During respiratory seasons, sensitivity reached 81% for SARS-CoV-2 and 81% for influenza A; both had 100% sensitivity during peak case weeks. Notably, off-peak influenza detections occurred in hospital wastewater at all three hospitals in summer 2024 without corresponding hospital case detection, suggesting the presence of undiagnosed cases. These findings underscore the potential utility of hospital-based wastewater monitoring for tracking respiratory virus activity.

## Introduction

Community-based wastewater surveillance has been used for decades to monitor infectious diseases(1) and has re-emerged as a powerful tool during the COVID-19 pandemic. Recent studies demonstrated strong correlations between SARS-CoV-2 RNA concentrations and corresponding inpatient hospitalization rates(2), showed that wastewater detection preceded clinical reporting by several days,(3,4) and identified variants of concern weeks before clinical detection.(5) By 2022, the Center for Disease Control and Prevention’s (CDC) National Wastewater Surveillance System (NWSS) covered hundreds of jurisdictions nationwide, offering population-level insights that did not depend on individual clinical/laboratory-based testing, or at-home testing, which became particularly valuable as both declined in the later stages of the pandemic.(6) Building on this infrastructure, wastewater surveillance has since expanded beyond SARS-CoV-2 to include influenza A/B, RSV, metapneumovirus, and other pathogens, with wastewater concentrations of these pathogens shown to correlate with emergency department visits and to support detection before traditional clinical surveillance signals begin to rise.(7,8)

However, community wastewater surveillance does not necessarily provide more geographically localized situational awareness, and there is no clinical or epidemiologic information, which is also needed for decision-making. Surveillance of hospital wastewater(9) reflects contributions from a smaller, better defined population of inpatients, outpatients, healthcare workers, and visitors interacting with a specific facility. Facility-level wastewater surveillance approaches have been explored in hospitals, long-term care facilities, airports, and university settings, largely with a focus on SARS-CoV-2. A multicenter study done in Calgary, Canada found that hospital wastewater SARS-CoV-2 viral burden correlated with both increasing inpatient case counts and hospital-associated transmission.(10) Hospital wastewater monitoring over three years in a New York hospital showed that rising SARS-CoV-2 wastewater concentrations preceded increases in inpatient caseloads by up to one week.(11) Facility-level surveillance has also been explored in nursing facilities and university dormitories, though with relatively short study durations.(12,13) More recently, a pilot study at two California emergency departments detected SARS-CoV-2, influenza A, and RSV in hospital wastewater using composite samplers, which capture viral particles over an extended deployment period rather than at a single point in time, yielding patterns generally consistent with clinical case trends.(9) This study evaluates the sensitivity, specificity, and positive predictive value (PPV) of hospital wastewater surveillance during May 2024-February 2026 for detecting SARS-CoV-2, influenza A, and influenza B within a single healthcare system.

## Methods

### Study Design & Setting

This retrospective study was conducted from May 2024-April 2026 at three New York City Health+Hospitals (H+H) facilities. NYC H+H is the largest municipal health care delivery system in the United States, operating 11 acute care hospitals across all five boroughs. New York City H+H serves approximately 1.4 million patients annually, including uninsured patients, visitors, and many individuals experiencing homelessness who may not otherwise be captured by routine clinical surveillance.(14) The three study facilities (Hospitals A, B, and C) capture geographic and demographic diversity within the H+H system, serving communities with annual inpatient admissions ranging from 12-21K annually. The H+H patient population is predominantly Hispanic/Latinx (42.9%) and Black and African-American (31.0%), with over 30% of patients primarily Spanish-speaking.(14) As part of an ongoing Biosurveillance Pilot Program at these facilities, hospital wastewater has been sampled approximately weekly at each site since May 2022.(15)

### Wastewater Sample Collection

Composite samples were collected using a passive sampler deployed at each collection point to capture viral particles and RNA from hospital wastewater. For Hospital A, wastewater was accessed via the main sewage outflow pipe located in the basement. For Hospitals B and C, manholes outside the buildings were used. Each passive sampling device consists of four screw-capped perforated cylinders, each enclosing a sachet filled with activated carbon (Figure S1). The cylinders are connected using nylon strings, are free to move independent of each other, and are then tethered to handrails or manhole lids while deployed for a 24-hour period before being retrieved, except at Hospital A where the sampling period was approximately 12 hours due to staffing constraints. Following the sampling period, cylinders from each sampler were transferred into a buffer solution and transported on ice for processing in the lab. The target sampling frequency was weekly with adjustments made for holidays and staffing constraints. Study team members collected and transferred samples to the laboratory and stored at 4°C until they were processed, which generally was within 48 hours of retrieval, depending on staff availability.

### Pre-analytic Processing and RNA Extraction

Nucleic acid yields were not significantly different when samplers were processed immediately as opposed to a 24-or 48-hour delay (data not shown). Total nucleic acids were extracted using a direct nucleic acid extraction method with a modified protocol 5B of the Wizard^®^ Enviro Total Nucleic Acid Kit (Promega, A2991). The four sachets from each sampler were transferred into a 50 mL falcon tube containing 20 mL of cold buffer 2 and incubated at room temperature for 30 minutes with gentle shaking. After decanting buffer 2, the sachets were washed twice with buffer 1, then gently shaken in a mixture of 10 mL RNase-free water and 200 μl protease solution for 30 minutes. After removing the sachets, steps starting from step 3 of the Wizard^®^ Enviro Total Nucleic Acid Kit were followed to extract the total nucleic acids. At the final step, 40 μl of pre-heated nuclease-free water was used twice to elute the nucleic acids in a total volume of 80 μl. Extracted total nucleic acids were stored at –20 °C until quantification.

### RNA Quantification

Wastewater samples were analyzed via digital PCR for four viral targets: SARS-CoV-2, influenza A, influenza B, and pepper mild mottle virus (PMMoV). Digital PCR was performed using the QIAcuity One 5plex dPCR system (Qiagen, Hilden, Germany) with the QIAcuity One-step Advanced Probe kit and a GoTaq Enviro Wastewater FluA, FluB, SC2 multiplex assay (Promega, AM2170) for absolute quantification of all four viral targets.(16) The reaction mixture was transferred into a QIAcuity 26k 24-well Nanoplate (Qiagen) and loaded into the QIAcuity One 5plex system. Partition thresholds were set using QIAcuity software, with proper separation between positive and negative partitions confirmed in all controls (Table S3). The RT-dPCR reaction mix, cycling conditions, passive sampler total gene copy calculation, and the digital MIQE standards (Table S1) are detailed in the supplementary information. Each sample was run in duplicate with positive and negative controls. Positive controls consisted of manufacturer-supplied in vitro transcribed RNA fragments of FluA, FluB, SC2, and PMMoV at 4 × 10^6^ copies/μl, and a no-template control was included in each run. The dPCR Summary statistics are shown in Table S2.

Raw viral concentrations were expressed in gene copies per microliter. Viral concentrations were normalized to PMMoV (Figure S2) to account for variation in flow rates, human waste contributions, and wastewater dilution across the three hospital sites.(17) Normalization was performed by dividing each target’s concentration by the corresponding PMMoV concentration and multiplying by 10^6^, yielding normalized copies per million PMMoV copies. PMMoV-normalized concentrations were log10-transformed for analysis.

### Hospital Case Data

We used aggregate hospital case data from the NYC H+H electronic health record (EHR) for all three hospitals for the duration of the study period. Case data included the number PCR-confirmed COVID-19 and influenza cases combined across inpatient, outpatient, and emergency department settings. Available case data did not distinguish between influenza A and influenza B subtypes. Citywide COVID-19 and influenza case data were obtained from the NYC Health Department’s publicly available Respiratory Illness data repository (18) and plotted alongside hospital-level results to assess whether facility-level wastewater signals and case trends (10-day rolling average) correlated with broader community disease activity (Figures 2, 3A, 3B).

## Statistical Analysis

We computed the sensitivity, specificity, and PPV of detecting SARS-CoV-2 and influenza A/B inpatient caseloads from wastewater using clinical cases as the ‘gold standard’. Specifically, diagnosed cases were summed by week. Weeks were classified as ‘true positive’ if they exceeded each of three minimum weekly case thresholds evaluated (≥1, ≥5, and ≥10 cases per week) and were classified as ‘true negative’ otherwise. Sensitivity was defined as the probability of detecting any level of a virus in wastewater during true positive weeks. Specificity was defined as the probability of not detecting any virus in wastewater during true negative weeks. PPV was defined as the probability that a week when virus was detected in wastewater coincided with a true positive week. Weeks in which no wastewater sample was collected were excluded from the analysis.

### Wave periods (surges)

Pathogen-specific wave periods were defined by visual inspection of combined weekly hospital case trends across all three facilities. The baseline for each pathogen was defined as the typical range of combined weekly case counts observed during periods of minimal case activity. Wave onset was identified as the first week in which combined case counts began a sustained increase over at least two to three consecutive weeks above this baseline range, and wave offset was defined as the point at which cases had declined to near-baseline levels. Wave period dates are reported in the footnotes for Table 3. Estimates are reported over three windows: the full study period (May 2024-April 2026), pathogen-specific hospital case wave periods, and the three peak weeks within each wave period.

### Ethical review

This work was designated as non-human subjects research by the IRB of the City University of New York (CUNY).

## Results

From May 2024 to March 2026, a total of 195 wastewater samples were collected and 190 were analyzed by digital polymerase chain reaction (dPCR) across the three facilities (Table 1 and Figure 1). SARS-CoV-2 was detected in 38.6%, 31.8%, and 38.8% of samples analyzed at Hospitals A, B, and C, respectively. Influenza A detection rates were 19.3%, 28.8%, and 31.3%, and influenza B detection rates were 12.3%, 16.7%, and 16.4%, respectively. Peak weekly COVID-19 case counts occurred during summer of 2024, with 64 cases at Hospital A (week of July 14, 2024), 43 cases at Hospital B (week of June 16, 2024), and 57 cases at Hospital C (week of June 30, 2024). Peak weekly influenza case counts were 186, 192, and 185 at Hospitals A, B, and C respectively, all occurring during the week of December 14, 2025, representingn approximately three times the peak weekly COVID-19 caseload. A comparison of the wastewater data, individual hospital clinical data, and citywide clinical data is shown in Figure 2. In general, hospital wastewater showed higher activity during wave peaks, but there were also off-peak periods where viruses were detected in wastewater at levels comparable to peak periods (Figure 2).

**Figure 1:**
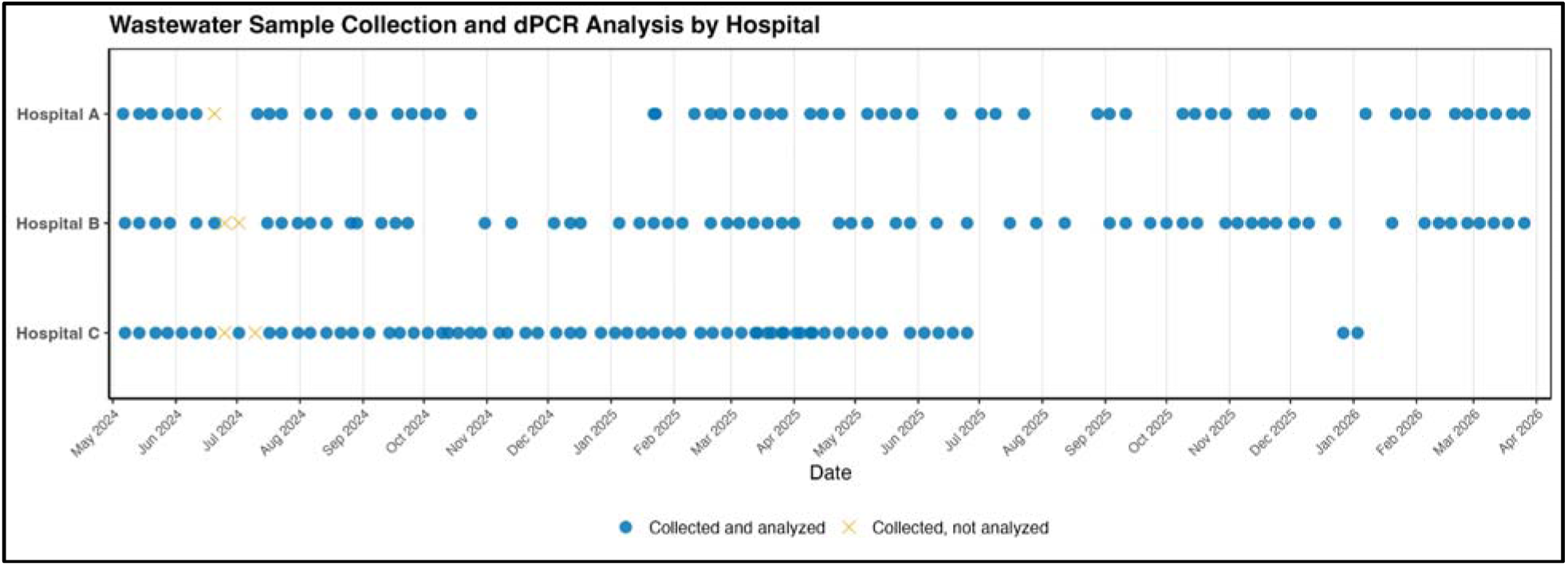
Wastewater Sample Collection and dPCR Analysis by Hospital from May 2024-April 2026. *Dates on the x-axis correspond to the dates the sample was collected.

**Figure 2:**
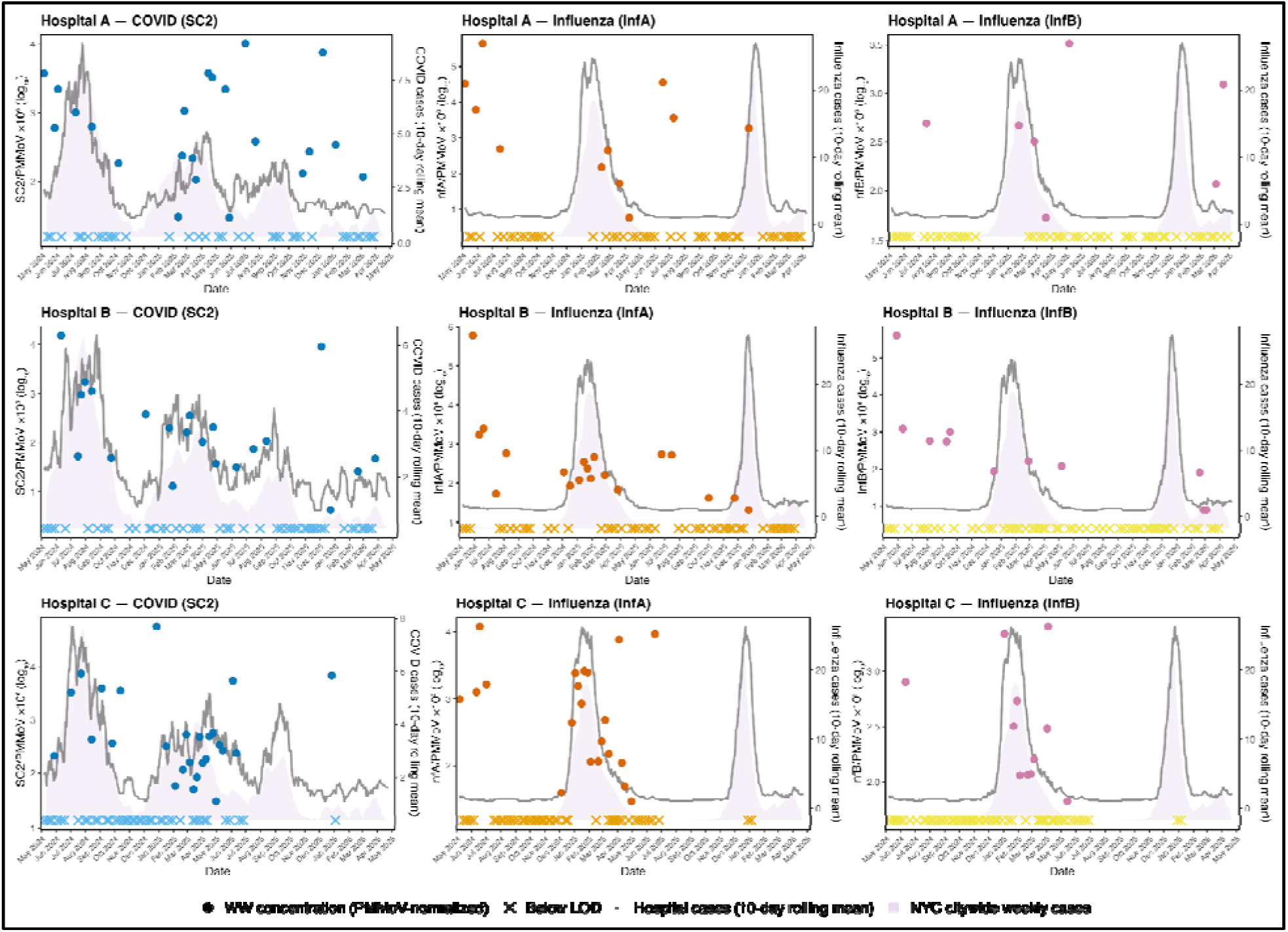
Wastewater Total Viral Particles vs. Hospital and NYC Citywide Case Trends by Pathogen, May 2024-April 2026. *Colored dots represent PMMoV-normalized wastewater viral concentrations (log_10_) from all three hospital sampling sites. X marks indicate samples below the limit of detection. The grey line represents the 10-day rolling mean of combined clinical case counts for each hospital. The pink shaded area represents NYC citywide weekly case counts (source: NYC DOHMH Respiratory Illness Data).

**Table 1:**
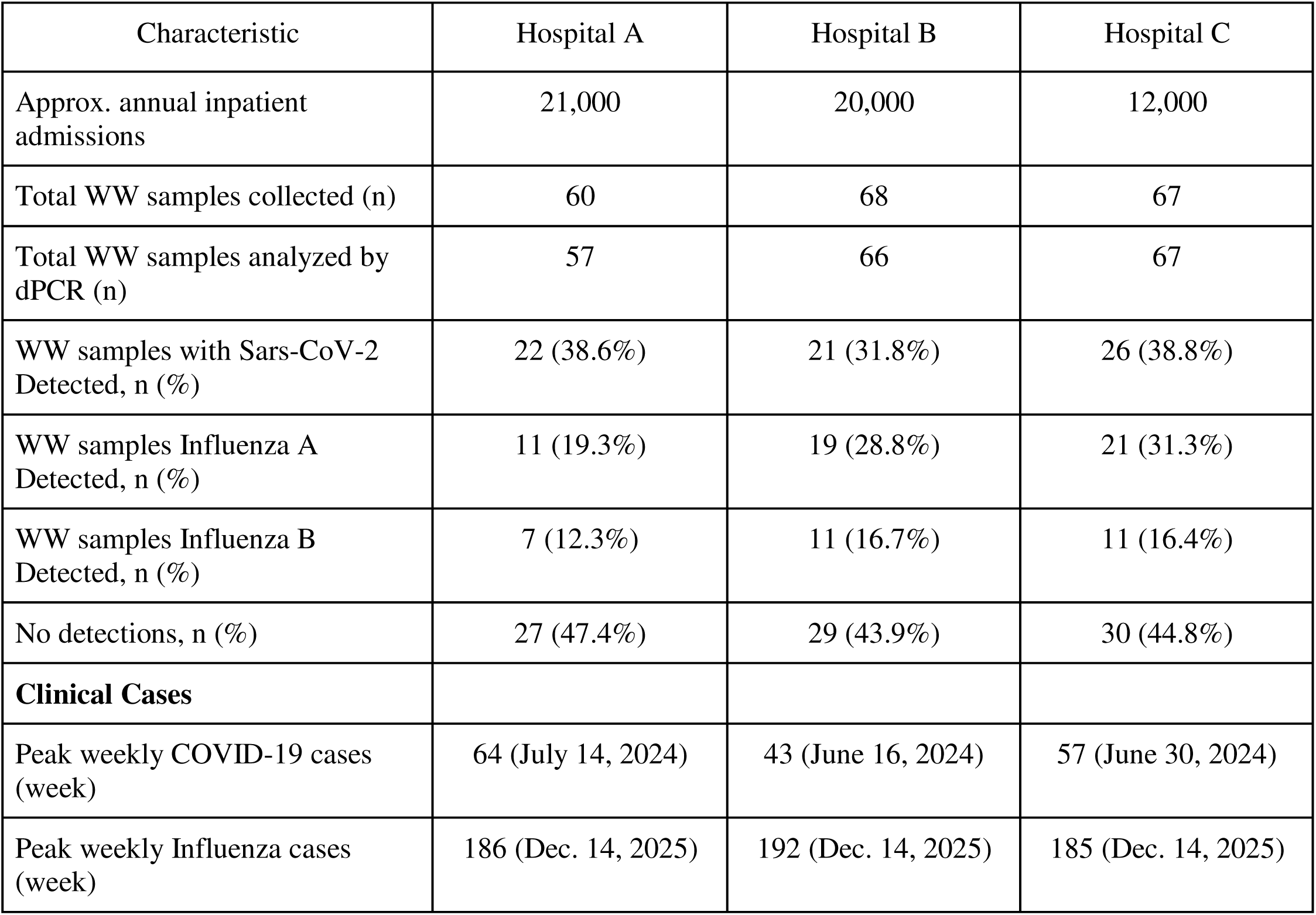
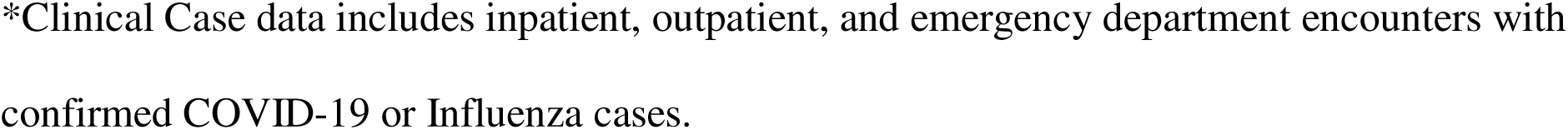
Descriptive Characteristics of Wastewater Sample collection and, and Clinical Cases by hospital.

| Characteristic | Hospital A | Hospital B | Hospital C |
| --- | --- | --- | --- |
| Approx. annual inpatient admissions | 21,000 | 20,000 | 12,000 |
| Total WW samples collected (n) | 60 | 68 | 67 |
| Total WW samples analyzed by dPCR (n) | 57 | 66 | 67 |
| WW samples with Sars-CoV-2 Detected, n (%) | 22 (38.6%) | 21 (31.8%) | 26 (38.8%) |
| WW samples Influenza A Detected, n (%) | 11 (19.3%) | 19 (28.8%) | 21 (31.3%) |
| WW samples Influenza B Detected, n (%) | 7 (12.3%) | 11 (16.7%) | 11 (16.4%) |
| No detections, n (%) | 27 (47.4%) | 29 (43.9%) | 30 (44.8%) |
| **Clinical Cases** |  |  |  |
| Peak weekly COVID-19 cases (week) | 64 (July 14, 2024) | 43 (June 16, 2024) | 57 (June 30, 2024) |
| Peak weekly Influenza cases (week) | 186 (Dec. 14, 2025) | 192 (Dec. 14, 2025) | 185 (Dec. 14, 2025) |

\*Clinical Case data includes inpatient, outpatient, and emergency department encounters with confirmed COVID-19 or Influenza cases.

### Sensitivity and Specificity of Wastewater Surveillance Across Case Thresholds

When data were combined across hospitals over the entire study period, sensitivity to detect a minimum of one case per week was 38% (95%CI: 31-46%) for SARS-CoV-2 and 36% (95%CI: 29-44%) for influenza (Table 2). Sensitivity increased at higher thresholds of the minimum number of cases shedding in hospital wastewater in a given week. For influenza, sensitivity of wastewater surveillance combined across hospitals rose from 36% (95%CI: 29-44%) to detect a minimum of 1 case to 46% (95%CI: 36-57%) to detect a minimum of 5 cases and 49% (95%CI: 37-61%) to detect a minimum of 10 cases within a week. For SARS-CoV-2, sensitivity combined across hospitals was 40% (95%CI: 32-48%) at ≥5 cases and 42% (95%CI: 34-52%) at ≥10 cases. The specificity could not be calculated for any pathogen at the ≥1 case threshold because all sampled weeks had at least one hospital case. For all three pathogens, PPV was 100% to detect at least one case, and remained high for SARS-CoV-2 to detect ≥5 cases (94%) and ≥10 cases (78%) per week. PPV for wastewater surveillance to detect influenza A or B was 72% (95%CI: 59-82%) at ≥5 cases and 60% (95%CI: 47-72%) at ≥10 cases per week.

**Table 2:** Sensitivity, Specificity, and positive predictive value of Hospital Wastewater Surveillance for Detecting SARS-CoV-2 and Influenza by Minimum Weekly Case Count Threshold, May 2024-April 2026.

**Threshold, May 2024- April 2026**
| Hospital | Target | Min. # cases detected | Sens-itivity | 95% CI | Spec-ificity | 95% CI | Positive Predictive Value | PPV 95% CI |
| --- | --- | --- | --- | --- | --- | --- | --- | --- |
| Hospital A | Flu A or B | ≥1 | 26% | 0.15, 0.40 | -n/d | - | 100% | 0.77, 1.00 |
| Hospital B | Flu A or B | ≥1 | 41% | 0.28, 0.54 | -n/d | - | 100% | 0.86, 1.00 |
| Hospital C | Flu A or B | ≥1 | 40% | 0.27, 0.54 | -n/d | - | 100% | 0.85, 1.00 |
| Combined | Flu A or B | ≥1 | 36% | 0.29, 0.44 | -n/d | - | 100% | 0.94, 1.00 |
| Hospital A | Flu A or B | ≥5 | 38% | 0.21, 0.56 | 90% | 0.70, 0.99 | 86% | 0.57, 0.98 |
| Hospital B | Flu A or B | ≥5 | 41% | 0.25, 0.59 | 60% | 0.39, 0.79 | 58% | 0.37, 0.78 |
| Hospital C | Flu A or B | ≥5 | 63% | 0.42, 0.81 | 82% | 0.63, 0.94 | 77% | 0.55, 0.92 |
| Combined | Flu A or B | ≥5 | 46% | 0.36, 0.57 | 77% | 0.66, 0.86 | 72% | 0.59, 0.82 |
| Hospital A | Flu A or B | ≥10 | 38% | 0.18, 0.62 | 81% | 0.64, 0.93 | 57% | 0.29, 0.82 |
| Hospital B | Flu A or B | ≥10 | 48% | 0.29, 0.68 | 67% | 0.47, 0.83 | 58% | 0.37, 0.78 |
| Hospital C | Flu A or B | ≥10 | 61% | 0.39, 0.80 | 75% | 0.57, 0.89 | 64% | 0.41, 0.83 |
| Combined | Flu A or B | ≥10 | 49% | 0.37, 0.61 | 74% | 0.64, 0.83 | 60% | 0.47, 0.72 |
| Hospital A | Sars-CoV-2 | ≥1 | 39% | 0.26, 0.52 | -n/d | - | 100% | 0.85, 1.00 |
| Hospital B | Sars-CoV-2 | ≥1 | 33% | 0.22, 0.46 | -n/d | - | 100% | 0.84, 1.00 |
| Hospital C | Sars-CoV-2 | ≥1 | 43% | 0.31, 0.57 | -n/d | - | 100% | 0.87, 1.00 |
| Combined | Sars-CoV-2 | ≥1 | 38% | 0.31, 0.46 | -n/d | - | 100% | 0.95, 1.00 |
| Hospital A | Sars-CoV-2 | ≥5 | 39% | 0.26, 0.54 | 67% | 0.22, 0.96 | 91% | 0.71, 0.99 |
| Hospital B | Sars-CoV-2 | ≥5 | 36% | 0.24, 0.50 | 100% | 0.54, 1.00 | 100% | 0.84, 1.00 |
| Hospital C | Sars-CoV-2 | ≥5 | 44% | 0.31, 0.59 | 67% | 0.22, 0.96 | 92% | 0.75, 0.99 |
| Combined | Sars-CoV-2 | ≥5 | 40% | 0.32, 0.48 | 78% | 0.52, 0.94 | 94% | 0.86, 0.98 |
| Hospital A | Sars-CoV-2 | ≥10 | 40% | 0.24, 0.57 | 63% | 0.38, 0.84 | 68% | 0.45, 0.86 |
| Hospital B | Sars-CoV-2 | ≥10 | 38% | 0.23, 0.54 | 75% | 0.53, 0.90 | 71% | 0.48, 0.89 |
| Hospital C | Sars-CoV-2 | ≥10 | 49% | 0.34, 0.64 | 82% | 0.48, 0.98 | 92% | 0.75, 0.99 |
| Combined | Sars-CoV-2 | ≥10 | 42% | 0.34, 0.52 | 72% | 0.58, 0.83 | 78% | 0.67, 0.87 |

\*n/d: Specificity could not be calculated at the ≥1 case threshold because all sampled weeks had at least one clinical case reported, resulting in no case-negative weeks for comparison.

**Table 3:**
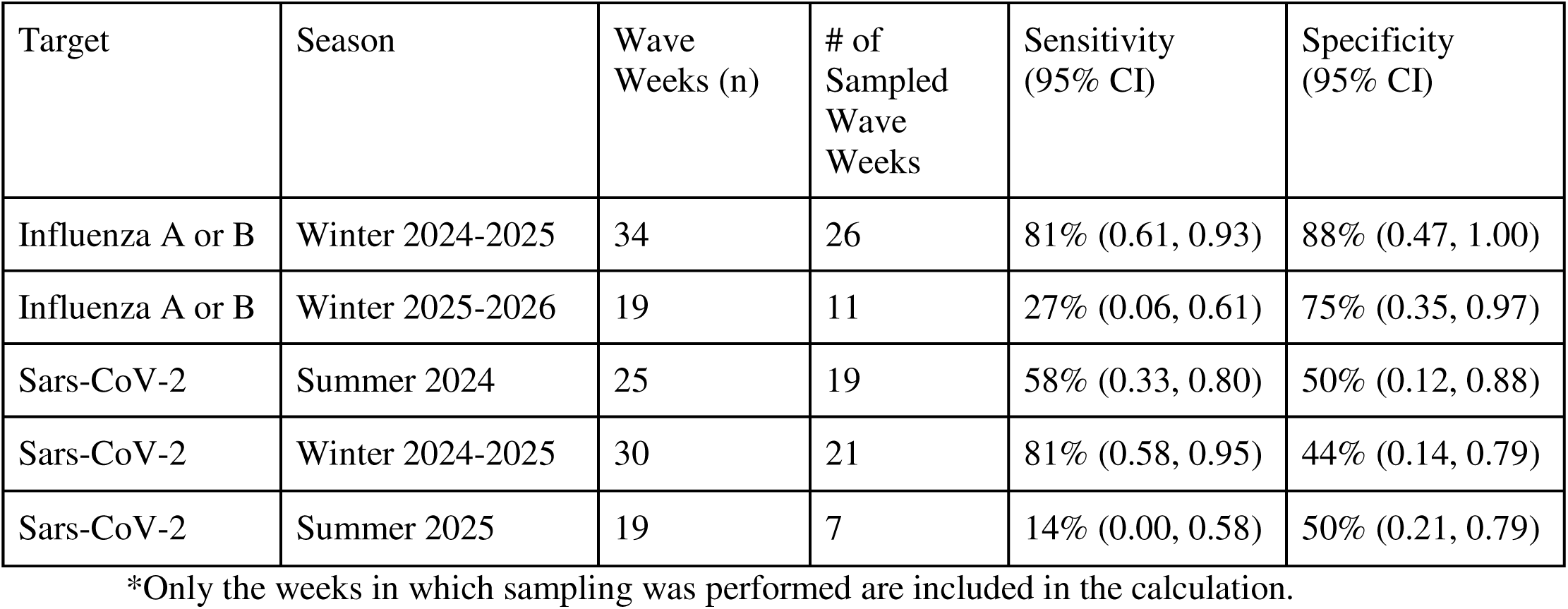
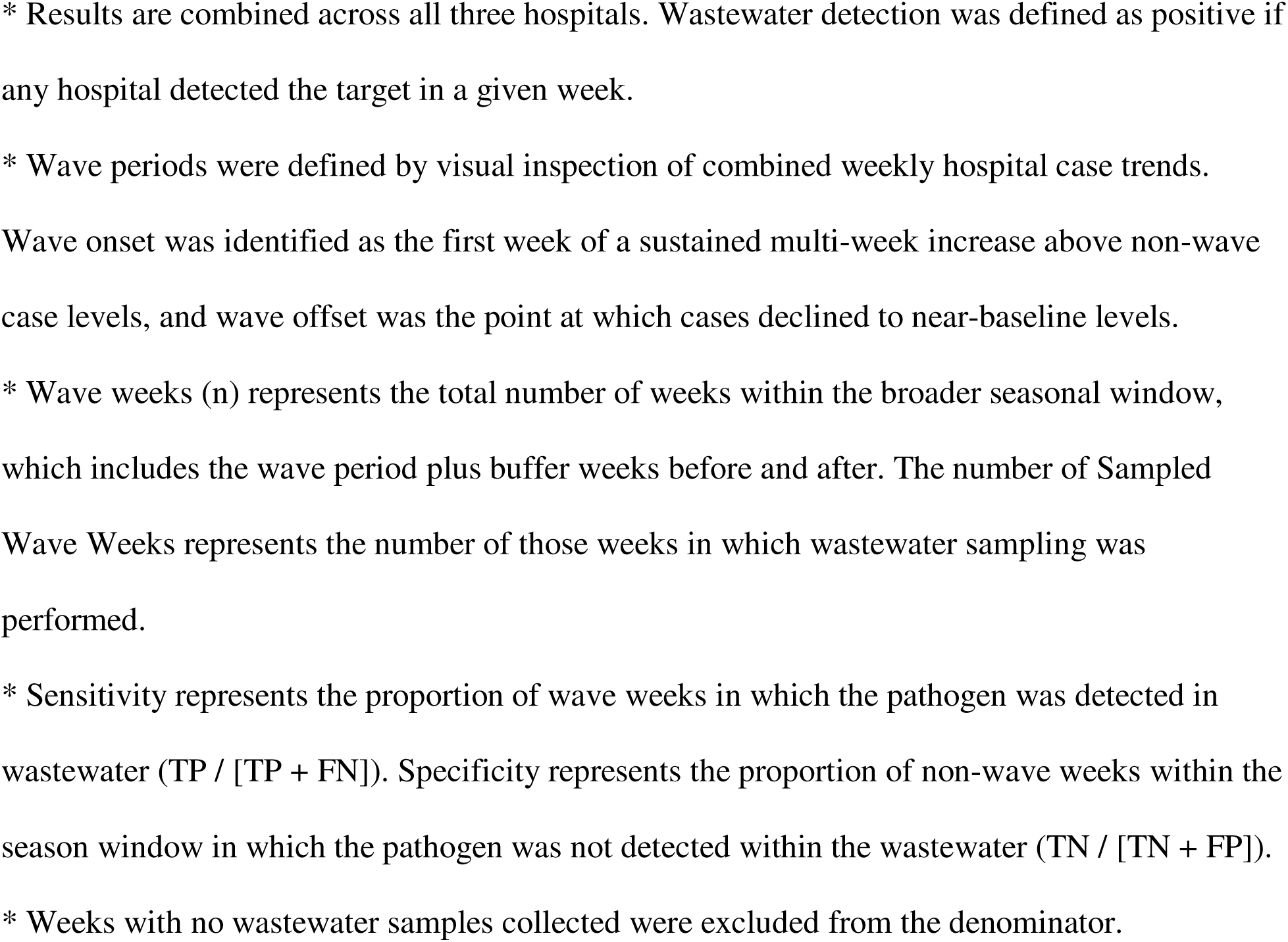
Sensitivity and Specificity of Hospital Wastewater Surveillance for Detecting SARS-CoV-2 and Influenza during Wave Periods, May 2024 - April 2026.

| Target | Season | Wave Weeks (n) | # of Sampled Wave Weeks | Sensitivity (95% CI) | Specificity (95% CI) |
| --- | --- | --- | --- | --- | --- |
| Influenza A or B | Winter 2024-2025 | 34 | 26 | 81% (0.61, 0.93) | 88% (0.47, 1.00) |
| Influenza A or B | Winter 2025-2026 | 19 | 11 | 27% (0.06, 0.61) | 75% (0.35, 0.97) |
| Sars-CoV-2 | Summer 2024 | 25 | 19 | 58% (0.33, 0.80) | 50% (0.12, 0.88) |
| Sars-CoV-2 | Winter 2024-2025 | 30 | 21 | 81% (0.58, 0.95) | 44% (0.14, 0.79) |
| Sars-CoV-2 | Summer 2025 | 19 | 7 | 14% (0.00, 0.58) | 50% (0.21, 0.79) |

\*Only the weeks in which sampling was performed are included in the calculation.
\* Results are combined across all three hospitals. Wastewater detection was defined as positive if any hospital detected the target in a given week.
\* Wave periods were defined by visual inspection of combined weekly hospital case trends.
Wave onset was identified as the first week of a sustained multi-week increase above non-wave case levels, and wave offset was the point at which cases declined to near-baseline levels.
\* Wave weeks (n) represents the total number of weeks within the broader seasonal window, which includes the wave period plus buffer weeks before and after. The number of Sampled Wave Weeks represents the number of those weeks in which wastewater sampling was performed.
\* Sensitivity represents the proportion of wave weeks in which the pathogen was detected in wastewater ( $TP / [TP + FN]$ ). Specificity represents the proportion of non-wave weeks within the season window in which the pathogen was not detected within the wastewater ( $TN / [TN + FP]$ ).
\* Weeks with no wastewater samples collected were excluded from the denominator.

**Appendix Table A:** Detailed Classification Counts for Wave Detection (Table 3)

| Target | Season | Wave Weeks (n) | # of Sampled Wave Weeks | # Weeks WW+ / Cases+ (TP) | # Weeks WW+ / Cases- (FP) | # Weeks WW- / Cases- (TN) | # Weeks WW- / Cases+ (FN) |
| --- | --- | --- | --- | --- | --- | --- | --- |
| Inf A or B | Winter 2024-25 | 34 | 26 | 21 | 1 | 7 | 5 |
| Inf A or B | Winter 2025-26 | 19 | 11 | 3 | 2 | 6 | 8 |
| SC2 | Summer 2024 | 25 | 19 | 11 | 3 | 3 | 8 |
| SC2 | Winter 2024-25 | 30 | 21 | 17 | 5 | 4 | 4 |
| SC2 | Summer 2025 | 19 | 7 | 1 | 6 | 6 | 6 |

\*This table provides the detailed classification counts that underlie Table 3 in the main text.

### Sensitivity and Specificity of Wastewater Surveillance by Seasonal Wave

The study period covered two winter waves for influenza A/B (Winter 2024-25 and Winter 2025-2026) and three waves for SARS-CoV-2 (Summer 2024, Winter 2024-25, and Summer 2025). No SARS-CoV-2 wave was observed during Winter 2025-26. Each wave was assessed separately for each pathogen, with results reported combined across all three hospitals. During the Winter 2024-25 influenza wave, wastewater sensitivity for influenza was 81% (95%CI: 61-93%) with a specificity of 88% (95%CI: 47-100%) (Table 3). During the Winter 2025-26 influenza wave, sensitivity was 27% (95%CI: 6-61%) with a specificity of 75% (95%CI: 35-97%), reflecting limited sampling during that period. For SARS-CoV-2, sensitivity was the highest during the Winter 2024-25 wave at 81% (95%CI: 58-95%) and 58% (95%CI: 33-80%) during the Summer 2024 wave. The Summer 2025 SARS-CoV-2 wave had a sensitivity of 14% (95%CI: 0-58%).

### Wastewater Detection During Peak Case Burden Weeks

Influenza case peaks were assessed for Winter 2024-25 and Winter 2025-26, and SARS-CoV-2 peaks were assessed for Summer 2024 and Winter 2024-25 separately. SARS-CoV-2 peaks were not assessed for Summer 2025 due to limited wastewater sampling or for Winter 2025-26, during which combined weekly case counts remained low and did not meet the criteria for a wave period. Influenza A or B was detected in wastewater during all three highest case burden weeks during Winter 2024-25 (3/3, 100%) and during 2 of 3 peak weeks during Winter 2025-26 (67%). SARS-CoV-2 was detected during all three highest case burden weeks during both Summer 2024 (3/3, 100%) and Winter 2024-25 (3/3, 100%) (Table 4).

**Table 4:**
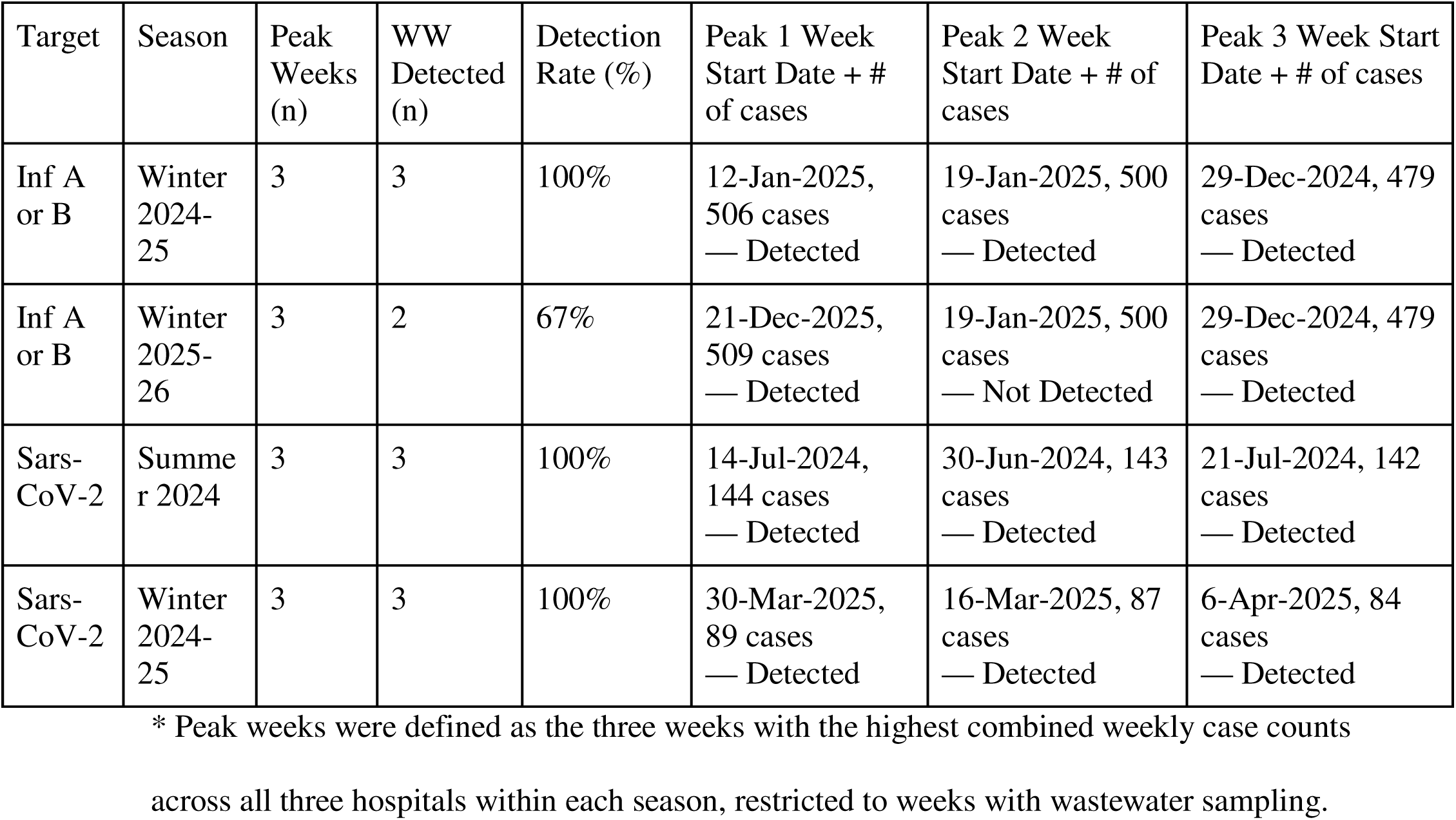
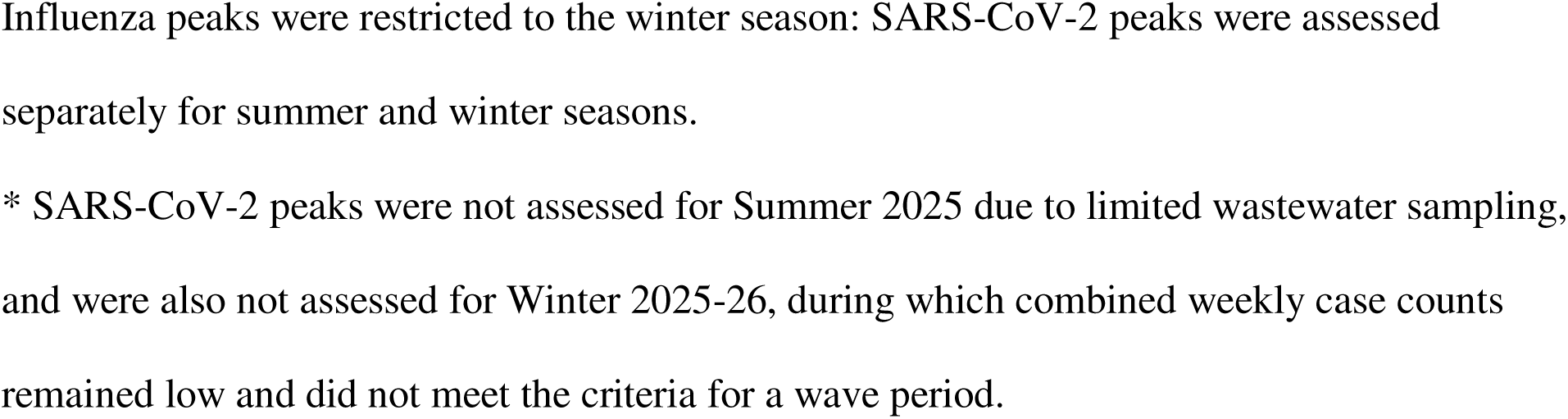
Wastewater Detection during the Three Highest Case activity weeks by Pathogen, among weeks with wastewater sampling, May 2024-April 2026.

| Target | Season | Peak Weeks (n) | WW Detected (n) | Detection Rate (%) | Peak 1 Week Start Date + # of cases | Peak 2 Week Start Date + # of cases | Peak 3 Week Start Date + # of cases |
| --- | --- | --- | --- | --- | --- | --- | --- |
| Inf A or B | Winter 2024-25 | 3 | 3 | 100% | 12-Jan-2025, 506 cases<br>— Detected | 19-Jan-2025, 500 cases<br>— Detected | 29-Dec-2024, 479 cases<br>— Detected |
| Inf A or B | Winter 2025-26 | 3 | 2 | 67% | 21-Dec-2025, 509 cases<br>— Detected | 19-Jan-2025, 500 cases<br>— Not Detected | 29-Dec-2024, 479 cases<br>— Detected |
| Sars-CoV-2 | Summer 2024 | 3 | 3 | 100% | 14-Jul-2024, 144 cases<br>— Detected | 30-Jun-2024, 143 cases<br>— Detected | 21-Jul-2024, 142 cases<br>— Detected |
| Sars-CoV-2 | Winter 2024-25 | 3 | 3 | 100% | 30-Mar-2025, 89 cases<br>— Detected | 16-Mar-2025, 87 cases<br>— Detected | 6-Apr-2025, 84 cases<br>— Detected |

\* Peak weeks were defined as the three weeks with the highest combined weekly case counts
across all three hospitals within each season, restricted to weeks with wastewater sampling.
Influenza peaks were restricted to the winter season: SARS-CoV-2 peaks were assessed separately for summer and winter seasons.
\* SARS-CoV-2 peaks were not assessed for Summer 2025 due to limited wastewater sampling, and were also not assessed for Winter 2025-26, during which combined weekly case counts remained low and did not meet the criteria for a wave period.

During summers of 2024 and 2025, influenza A was detected in wastewater at all three hospitals without corresponding increases in hospital influenza cases (Figures 2 and 3B). Influenza B was also detected sporadically during summer 2024, most frequently at Hospital B. During these periods, weekly influenza A/B case counts across the three hospitals averaged 1-3 cases per week, with multiple weeks reporting zero cases.

**Figure 3A:**
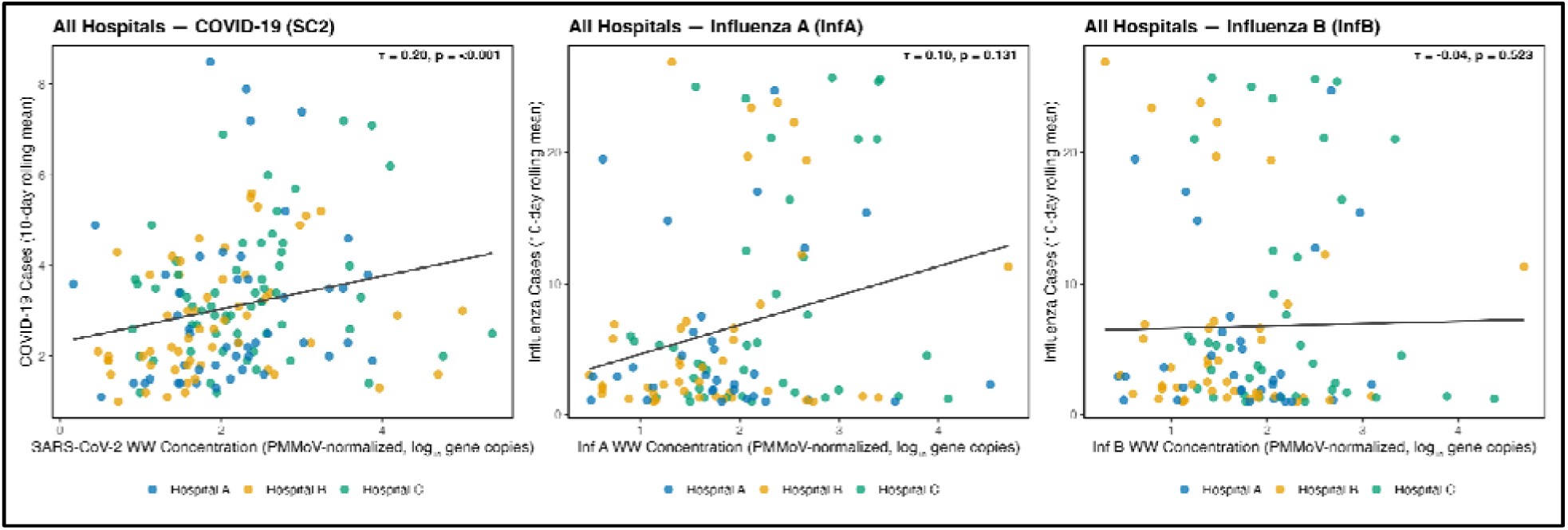
Scatter Plots of Wastewater Concentration vs. Case Counts by Pathogen (All Hospitals), May 2024 - April 2026. *Each point represents a single wastewater sampling event at one hospital site. The x-axis shows PMMoV-normalized wastewater viral concentration (log_10_) and the y-axis shows the corresponding 10-day rolling mean of hospital clinical case counts. The grey line represents the linear trend across all hospitals. Kendall’s tau and p-values are shown for the combined data across all three hospitals.

**Figure 3B:**
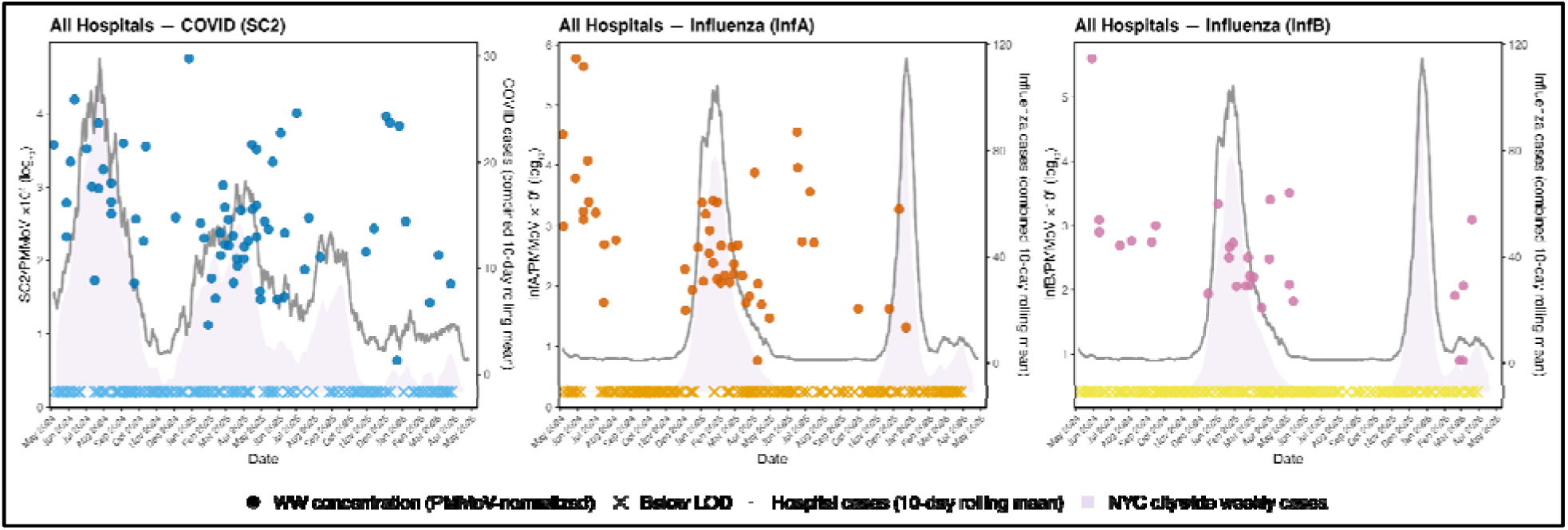
Combined Wastewater Viral Concentration vs. Hospital and NYC Citywide Case Trends by Pathogen (All Hospitals), May 2024 - April 2026. *Colored dots represent PMMoV-normalized wastewater viral concentrations (log_10_) from all three hospital sampling sites. X marks indicate samples below the limit of detection. The grey line represents the 10-day rolling mean of combined clinical case counts for each hospital. The pink shaded area represents NYC citywide weekly case counts (source: NYC DOHMH Respiratory Illness Data).

## Discussion

This study evaluated the ability of hospital-based wastewater surveillance to detect SARS-CoV-2, influenza A, and influenza B at three NYC H+H facilities over 23 months. When restricted to periods of increased case activity, the sensitivity of hospital wastewater to detect hospital cases during the Winter 2024-25 wave was high, reaching 81% for both SARS-CoV-2 (95%CI: 58-95%) and influenza (95%CI: 61-93%) when sampling coverage was highest. Sensitivity was lower during peak wave periods with fewer sampled weeks, including the Winter 2025-26 influenza wave (27%, 11 sampled weeks) and the Summer 2025 SARS-CoV-2 wave (14%, 7 sampled weeks). Specificity during peak wave periods was high for influenza A/B (75-88%) but more variable for SARS-CoV-2 (44-50%), and PPV ranged from 78-100% for SARS-CoV-2 and 60-100% for influenza A/B. SARS-CoV-2 and influenza A/B were both detected in wastewater during all three highest case burden weeks during Winter 2024-25.

Our findings suggest that wastewater surveillance and clinical surveillance capture different dimensions of respiratory virus activity within a healthcare facility. In particular, Influenza A was detected in hospital wastewater at all three facilities during summer 2024 without corresponding detection of any hospital cases, while there were also weeks when cases were diagnosed in the facility without corresponding wastewater detections. More generally, clinical data reflect those individuals who are tested and diagnosed, while wastewater captures viral shedding (over a 24-hour period in our study) from the full population present in the facility, including staff, visitors, and patients who are not tested. Capturing wastewater samples representative of an entire week in a hospital setting is challenging and resource intensitve. However, a 24-hour sampling period captures daily variability in hospital sewage flow, visitor activity, staff shift changes, and the timing of patient admission. To address the logistical challenges of repeated single timepoint ‘grab sampling’, we used a proprietary solid binding matrix deployed for 24 hour increments as a passive sampler, which has been shown to capture signals that grab sampling misses (19) and to perform comparably to autosamplers for SARS-CoV-2 detection.(20)

Our findings are generally consistent with prior work, though most existing studies have focused exclusively on SARS-CoV-2. Prior hospital wastewater studies have demonstrated correlations between SARS-CoV-2 viral burden and inpatient case counts, with wastewater concentrations preceding clinical increases by up to one week at some facilities.(10,21,22) A pilot study at two California emergency departments also detected SARS-CoV-2, influenza A, and RSV using composite samplers 3x/week, with patterns generally consistent with clinical data.(9) They detected SARS-CoV-2 in 38-67% and influenza A in 32-37% of samples, comparable to our detection rates (32-39% for SARS-CoV-2, 19-31% for influenza A).(9)

Daily wastewater volume at hospital facilities varies with the number of people contributing to the wastewater stream, including inpatients, outpatients, staff, and visitors, which fluctuates day-to-day and week-to-week. To address this, we used PMMoV normalization to compare measurements across time and facilities, accounting for different population sizes and flow rates.(9,11,21,22) Beyond dilution, which affects measurements for all pathogens detected in wastewater surveillance, fecal shedding varies considerably by pathogen. SARS-CoV-2 RNA is shed in feces at high concentrations (23)(24). In contrast, fecal shedding of influenza viruses is more variable and generally is present in the stool at lower concentrations than SARS-CoV-2.(25) Additionally, peak respiratory viral shedding for influenza A occurs within the first 1-2 days of symptom onset and declines rapidly thereafter, while influenza B exhibits a more prolonged shedding pattern with an earlier peak that can begin up to 2 days before symptom onset.(26) These differences suggest that by the time many influenza patients present for hospital care, they may have already passed peak viral shedding and may contribute less virus to the hospital wastewater in comparison to COVID-19 patients, who continue to shed SARS-CoV-2 in feces throughout their hospitalization.(24) Our data reflect this pattern, with higher detection for SARS-CoV-2, followed by influenza, consistent with existing literature.(9)

The lower PPV for influenza at higher case thresholds may reflect wastewater detections during summer 2024 that occurred without corresponding facility case increases, though confidence intervals were wide and differences between pathogens should be interpreted cautiously. During summer of 2024, influenza A was detected in hospital wastewater at all three hospitals without corresponding detection of clinical influenza cases. The viral concentrations in wastewater may have corresponded to undiagnosed patients, staff or visitors during a time when influenza testing is less common and thus not captured in the clinical case counts. Indeed NYC DOHMH influenza surveillance data (18) showed that influenza A and B were circulating in the community during the same period (Figure 3B). Overall, these detections highlight that wastewater captures viral shedding from the full spectrum of individuals present in a facility, not only those who tested positive and were reflected in diagnosed cases. Comparison with NYC citywide case trends showed that case patterns at the hospitals were generally consistent with community-level activity, providing context for interpreting weeks where facility wastewater and hospital case data diverged.

Future efforts to assess the potential added utility and complementarity of hospital-based wastewater surveillance should prioritize increasing sampling frequency to improve comparisons to daily clinical data, as prior work has suggested that sampling at least three times weekly is needed to maintain significant correlations with clinical trends.(11) In our study, weekly sampling was insufficient to assess lead-lag relationships between wastewater and clinical case data. In addition, incorporating information on admission and discharge dates for patients who tested positive for influenza and COVID-19 would provide a more accurate measure reflecting the expected viral contribution to hospital wastewater on or around the day of sampling. However, we could not examine the data this way because we didn’t have access to the individual level clinical data.

This study has several strengths. We simultaneously evaluated longitudinal data on three respiratory pathogens within the same hospital system, which allows for direct comparison of wastewater detection across viruses with different shedding characteristics.The study period covered both summer and winter waves for COVID-19 and two influenza seasons. Our work was conducted in a hospital system serving diverse, urban populations, which enhances the relevance of our findings for facilities serving similar communities that were disproportionately affected by COVID-19 and other respiratory virus surges.

There are also important study limitations. While we sampled for 24 hours once per week, studies suggest 2-3 samples per week are needed for correlation analyses between wastewater and clinical data.(25,27,28) There were also sampling gaps during holiday periods that coincided with peak respiratory virus circulation, reducing the number of weeks that we could assess during critical time periods. There was also limited wastewater sampling during the Summer 2025 SARS-CoV-2 wave and during the Winter 2025-26 influenza season. Also, we were not able to fully leverage the EHR to provide added epidemiologic and clinical context to the wastewater signals.

In conclusion, hospital wastewater monitoring is reflective of clinical case trends, but also provides complementary information. In addition, it can enhance a healthcare facility’s capacity to detect respiratory virus activity across the full spectrum of patients, staff, and visitors, and can provide infection control insights that hospital case data alone may not be able to fully capture. Integrating hospital-based wastewater surveillance with community-based wastewater and clinical surveillance data can also broaden public health monitoring of infectious diseases for better situational awareness.

## Data Availability

All data produced in the present study are available upon reasonable request to the authors, conditional on available resources.

## Supplementary Information

**Figure S1.**
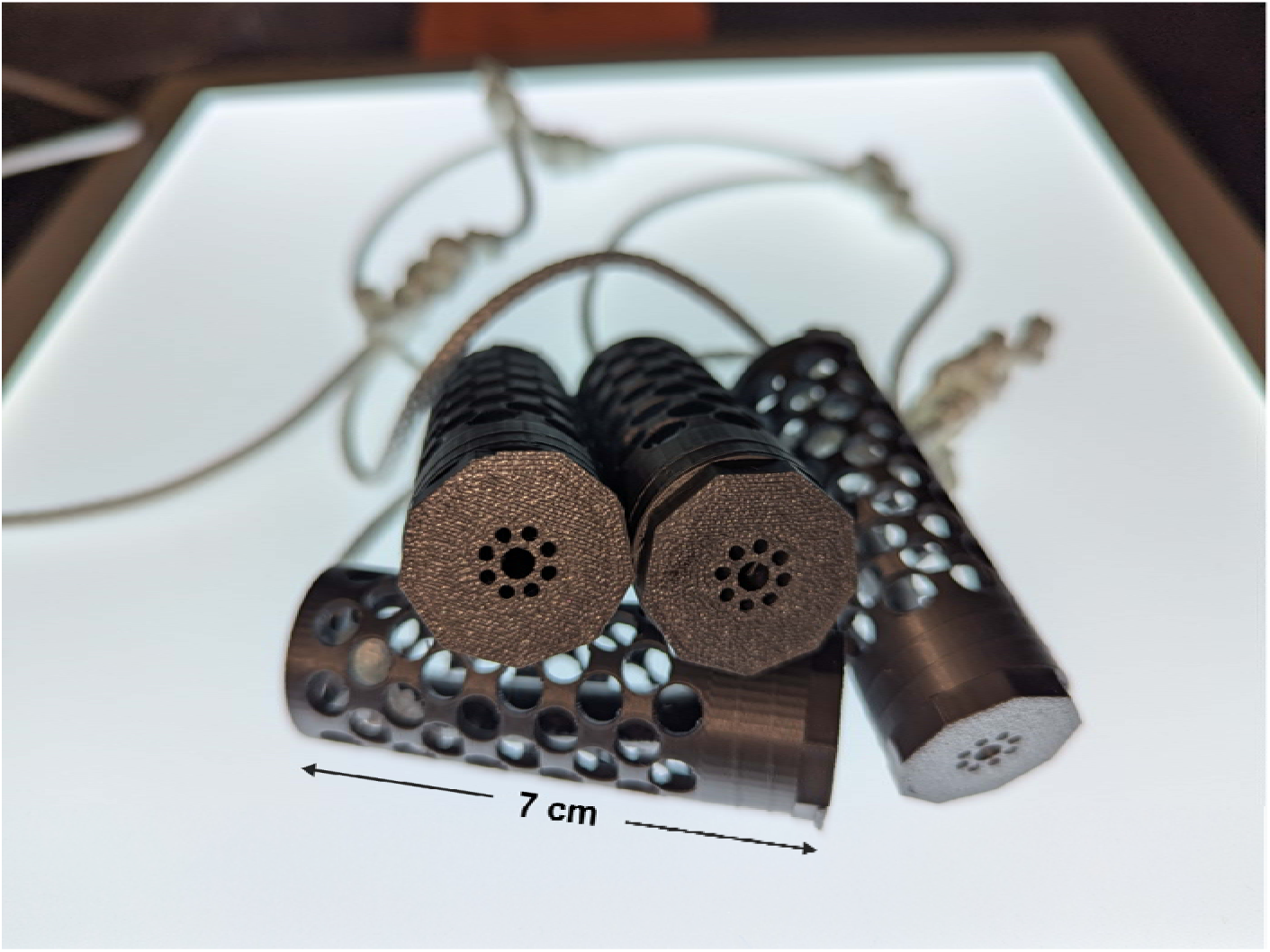
A passive sampler consisting of four screw-capped perforated cylinders, each housing a sachet filled with activated carbon. Each cylinder is fastened to a main nylon rope, which gives them mobility independent of each other. The main rope is tethered to handrails or manhole lids while deployed.

**Figure S2.**
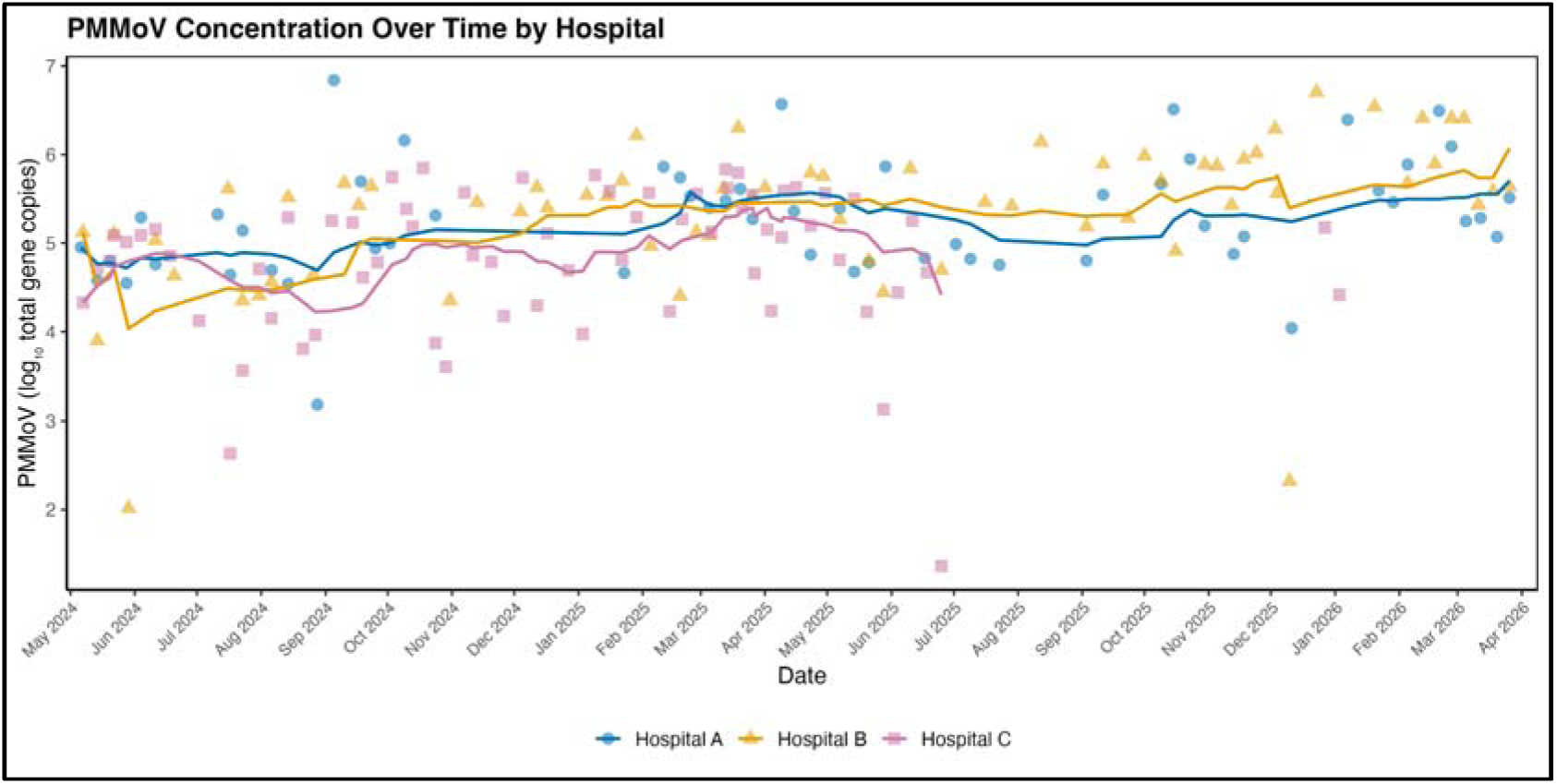
PMMoV Concentrations Across Three Hospitals, May 2024-April 2026

## RT-dPCR Reaction

The RT-dPCR reaction was prepared by mixing 10 _μ_l of OneStep (OS) Advanced Probe Master Mix, 0.4 _μ_l OS Advanced RT Mix, 2 _μ_l of FluA/FluB/SC2/PMMoV Primer/Probe Mix (Promega, AM2170), 4 _μ_l of sample, and molecular-grade water to a final volume of 40 _μ_l. InfA (FAM), InfB (HEX), SARS-CoV-2 (ROX), and PMMoV (Cy5) were measured in the same multiplex reaction with RT-dPCR reactions prepared as above. The cycling conditions included: reverse transcription, 50 °C for 40 min; 95 °C for 2 min; and 40 cycles of 95 °C for 5 s, 60 °C for 30 s.

## Passive sampler gene copy calculation (total copies extracted from each sampler)

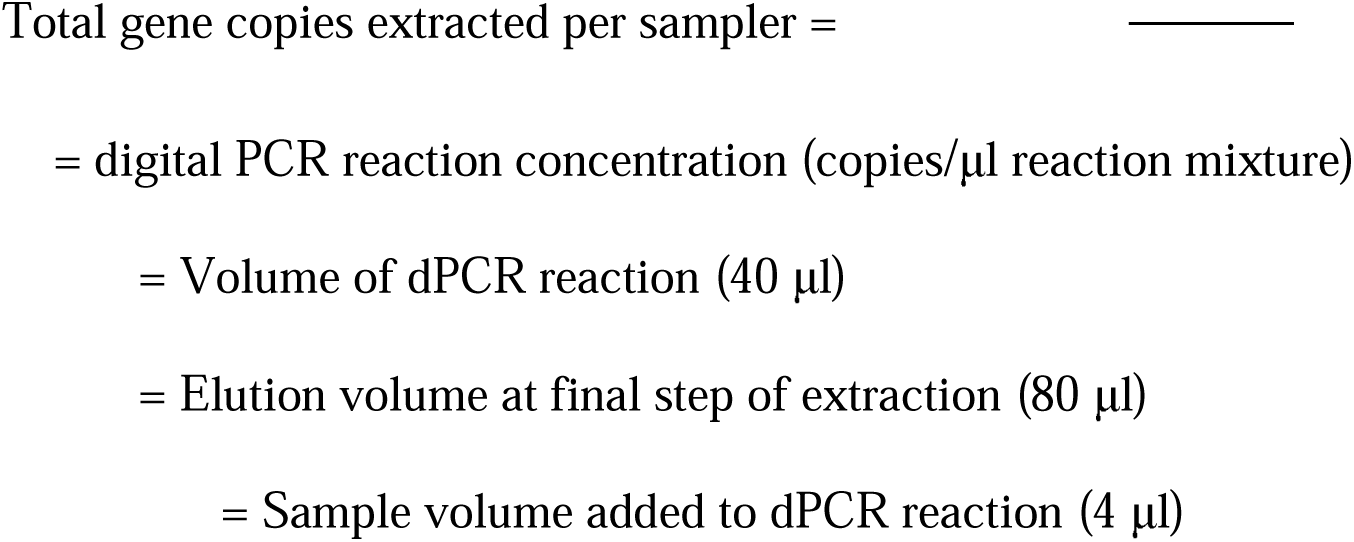

---

**Table S1.** Digital MIQE Checklist.

| ITEM TO CHECK | PROVIDED (Y/N) | COMMENT |
| --- | --- | --- |
| 1. SAMPLE | Y |  |
| Description | Y |  |
| Processing procedure | Y |  |
| Sample storage conditions and duration | Y |  |
| 2. NUCLEIC ACID EXTRACTION |  |  |
| Description of method |  |  |
| Solvent and volume used for elution |  |  |
| Storage conditions: temperature, concentration |  |  |
| 3. NUCLEIC ACID ASSESSMENT |  |  |
| Method evaluating nucleic acid quality | N |  |
| Method to quantify nucleic acid | N |  |
| Dilution steps to prepare working stocks | N | N/A |
| 4. NUCLEIC ACID MODIFICATION |  |  |
| Template modification | N | N/A |
| Re-purification following modification | N | N/A |
| 5. REVERSE TRANSCRIPTION |  |  |
| One or two step protocol | Y |  |
| Amount of nucleic acids per reaction | Y |  |
| Manufacturer of reagents and cat. no. | Y |  |
| Detailed reaction components and conditions | Y |  |
| cDNA priming method + concentration | N | N/A |
| Storage of cDNA: temperature, concentration | N | One step |
| 6. dPCR OLIGONUCLEOTIDE DESIGN AND TARGET INFORMATION |  |  |
| Primer/Probe sequences | N | Proprietary |
| Manufacturer details | Y |  |
| 7. dPCR PROTOCOL |  |  |
| Instrument manufacturer | Y |  |
| Complete thermocycling parameters | Y |  |
| Buffer/kit manufacturer and details | Y |  |
| Reaction volume and sample amount | Y |  |
| Primer/probe concentration | N | Proprietary |
| Template treatment (heating or chemical) | N | N/A |
| Polymerase identity and concentration, Mg<sup>++</sup> and dNTP concentrations | N | Qiagen Master mix |
| 8. dPCR VALIDATION |  |  |
| Optimization data for the assay | N | N/A |
| Analytical specificity and limit of blank (LOB) | N |  |
| Analytical sensitivity/LOD and method | N |  |
| Testing of inhibitors | Y |  |
| 9. DATA ANALYSIS |  |  |
| Mean copies per partition ($\lambda$) | Y | |
| dPCR analysis program (source, version) | Y |  |
| Results of positive and no-template controls | Y |  |
| Examples of positive experimental results | Y |  |
| Description of technical replication | Y |  |
| Average number of partitions measured | Y |  |
| Average petition volume | Y |  |
| Reproducibility (inter-experiment/user/lab etc.) | N |  |
| Description of normalization method | Y |  |
| Statistical method used for analysis | Y |  |

**Table S2.**
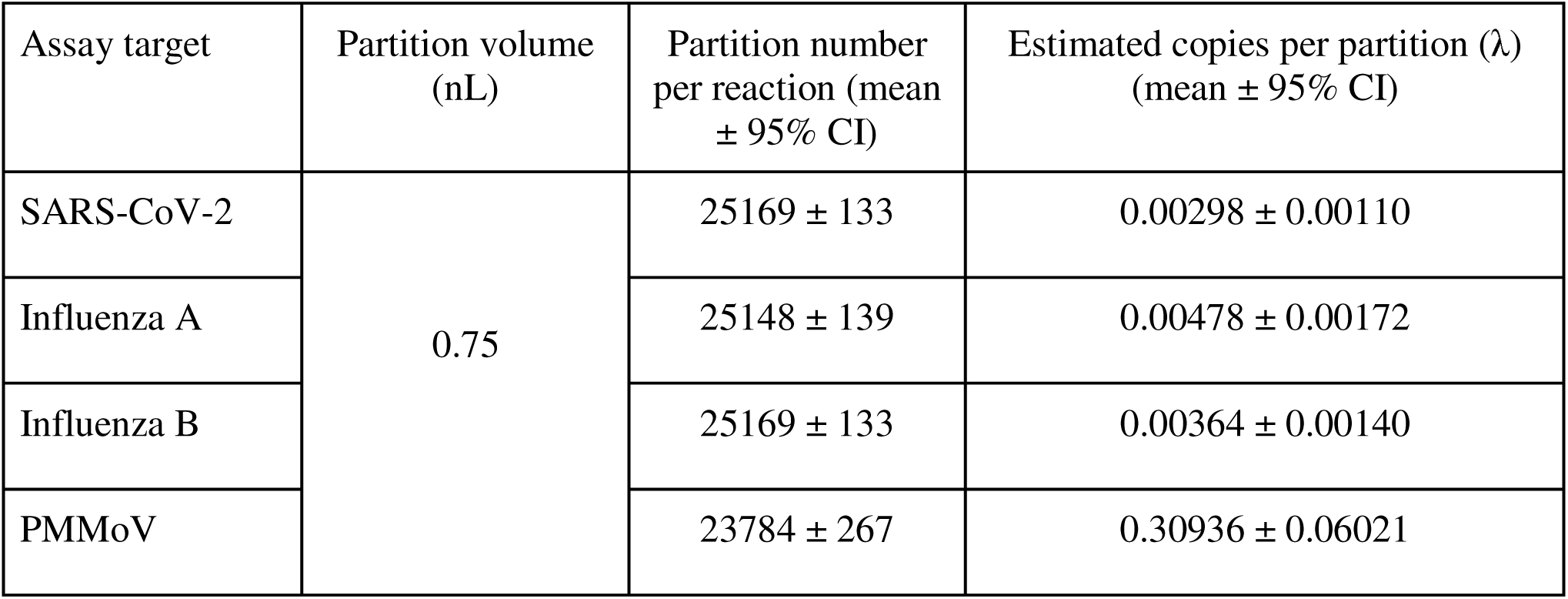
dPCR summary statistics.

**Table S3.**
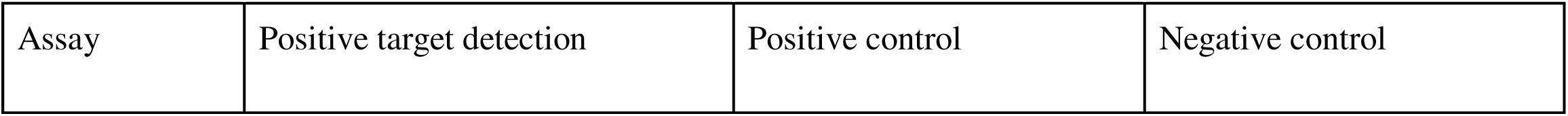
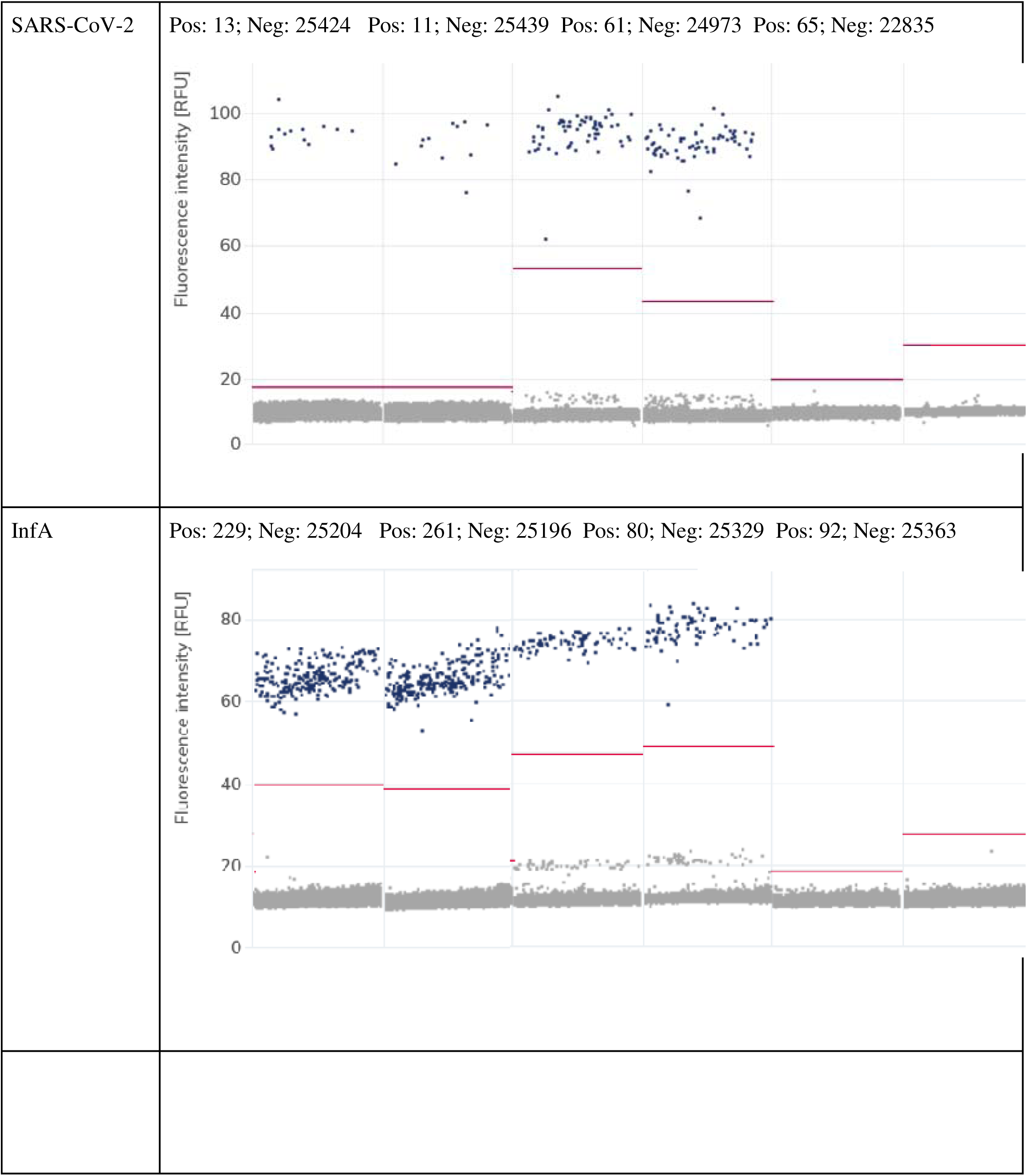
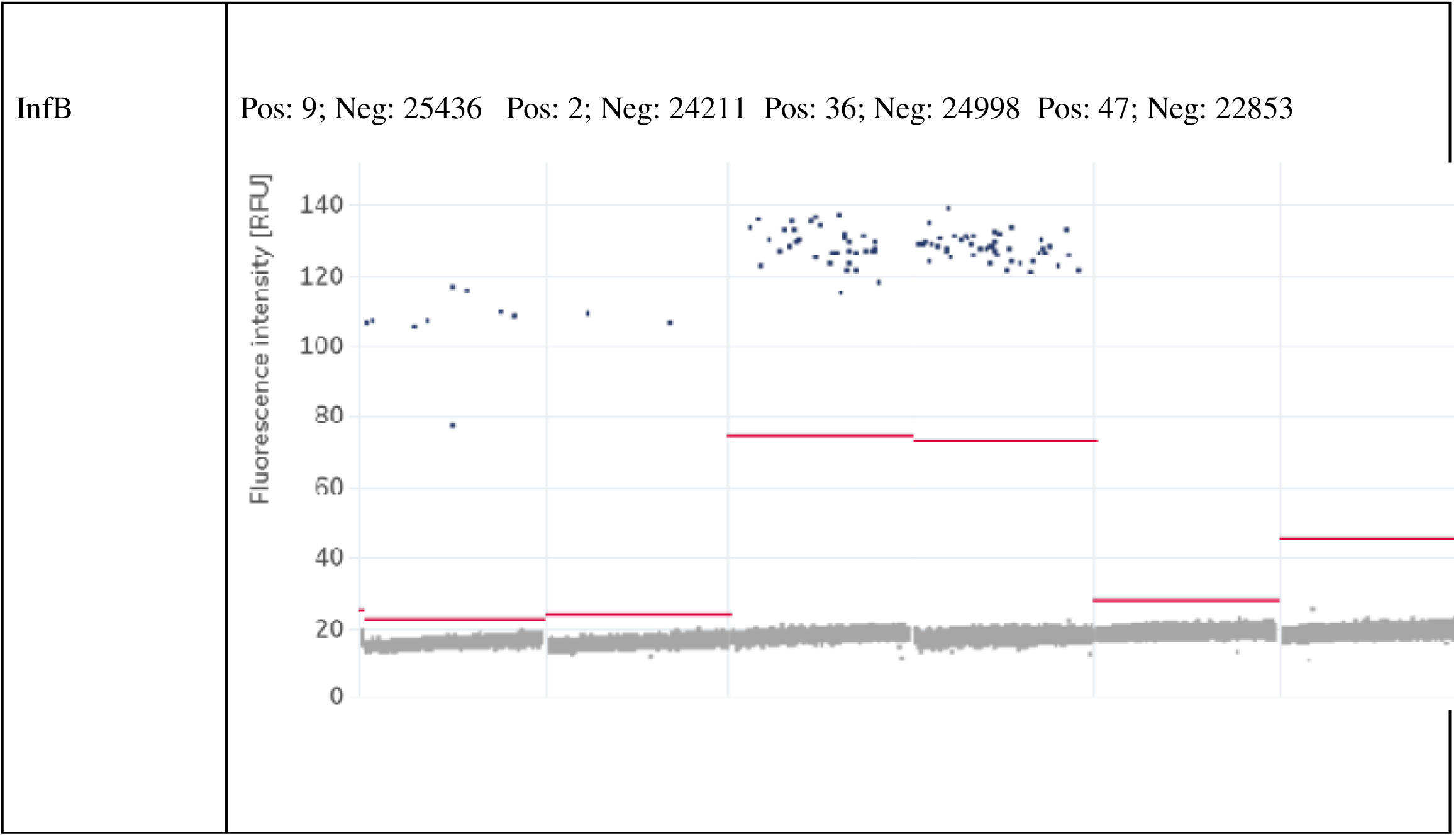
Examples of 2D scatterplots from dPCR output.

|  |  |  |  |
| --- | --- | --- | --- |
| Assay | Positive target detection | Positive control | Negative control |

## Acknowledgements

We thank the staff at the three NYC Health + Hospitals locations for their support in facilitating the wastewater sample collection. We thank Rebecca Posner for her assistance with sample extraction. Additionally, Rebecca and Kayla Wang assembled the devices used in this paper, we thank their help. We also thank the New York City Department of Health and Mental Hygiene for making citywide respiratory illness surveillance data publicly available for comparison. SP was supported by the Ferguson Emerging Infectious Diseases Fellowship of the Kennedy Krieger Institute.

